# Rates of oral nicotine pouch experimentation, initiation, and cessation by other tobacco product use and sociodemographic characteristics among U.S. adults, 2022–2024

**DOI:** 10.64898/2026.10.05.26364785

**Authors:** Olivia K. Roberts, Luz M. Sánchez-Romero, Evelyn Jimenez-Mendoza, Jihyoun Jeon, Nargiz Travis, Ritesh Mistry, Rafael Meza, David T. Levy, Andrew F. Brouwer

## Abstract

**Importance:** Oral nicotine pouches (ONPs) are tobacco-leaf-free nicotine products that have experienced a substantial increase in sales in recent years.

**Objective:** To determine how ONP experimentation, initiation, and cessation rates differ by other tobacco product use and sociodemographic characteristics.

**Design:** Nationally representative longitudinal cohort study; data from Waves 7 and 8 (2022–2024) of the Population Assessment of Tobacco and Health (PATH) Study.

**Setting:** US civilian, non-institutionalized population.

**Participants:** Adults in Waves 7 and 8 of PATH. Of 27,800 eligible participants, 24,640 with complete information on the exposure and outcome were included.

**Exposures:** Other tobacco product use (cigarette-only, electronic nicotine delivery system [ENDS]-only, smokeless tobacco [SLT]-only, and multiproduct use) and sociodemographic characteristics (sex, race and ethnicity, age, and educational attainment).

**Main Outcomes:** Hazard rates and ratios of transitioning between never, current, and non-current ONP use, defined by ever and past-30-day use, overall and by exposure group. Rates are estimated in univariable weighted multistate transition models.

**Results:** Among 24,640 participants, 23,555 (96.7%) never used, 802 (2.5%) non-currently used, and 283 (0.8%) currently used ONPs in 2022. From 2022–24, ONP initiation was higher among those using cigarettes only (HR 3.34, 95% CI 1.59–7.02), ENDS only (HR 9.68, 95% CI 2.89–32.5), SLT only (42.4, 95% CI 22.3–80.7), or multiple products (HR 18.4, 95% CI 7.11–47.8) compared to those who did not use these tobacco products. Those who used cigarettes only (HR 3.02, 95% CI 1.24–7.34), ENDS only (HR 2.03, 95% CI 1.11–3.71), or multiple products (HR 2.61, 95% CI 1.12–6.09) discontinued ONPs at higher rates than those using no other products. Men had higher ONP initiation and lower cessation rates than women, adults ages 35+ years had lower initiation rates than those ages 18–24, and non-Hispanic Black participants had higher cessation rates than non-Hispanic White participants.

**Conclusions and Relevance:** Initiation of ONPs was higher among those who used other tobacco products, highlighting the need to investigate whether ONPs replace these products or lead to poly-tobacco use. Higher cessation rates among those using cigarettes only or ENDS only may suggest experimentation rather than sustained use.

**Key Points:** *Question:* How do rates of oral nicotine pouch (ONP) experimentation, initiation, and cessation depend on other tobacco product use and sociodemographic characteristics?

*Findings:* Among 24,640 adults in a nationally representative longitudinal cohort study, individuals using other tobacco products experimented with and initiated ONPs at higher rates than those not using other tobacco products, but all except those using only smokeless tobacco also discontinued ONPs at higher rates.

*Meaning:* Adults using other tobacco products were more likely to experiment with but not necessarily continue to use ONPs in this period in which ONPs were recently introduced.

## Introduction

Oral nicotine pouches (ONPs) are synthetic or tobacco-derived nicotine products that have similar nicotine delivery levels as snus or moist snuff pouches.^1^ Initially introduced in 2016, sales of ONPs in the US increased substantially starting in 2019, especially among brands such as Zyn and On!.^2,3^ The potential health effects of ONP use are currently not well understood, but are likely to be substantially less than those of combustible tobacco.^4^ Parallel to the ongoing debate around the introduction of e-cigarettes and other electronic nicotine delivery systems (ENDS), debates center around whether ONPs could be utilized to aid cigarette cessation and whether ONPs lead to nicotine dependency among tobacco-naive individuals.^5,6^

Understanding how experimentation, initiation, and cessation of these products depend on other product use, as well as monitoring changes over time, will be necessary to project future ONP use, assess public health impacts, and develop interventions and regulations to reduce the risk of any potential harms, while maximizing any potential health gains.^7^ We adapted a multistate transition model, previously developed to assess rates of product transitions among cigarette, ENDS, and dual use,^8^ to estimate rates of short-lived experimentation, initiation, cessation, and relapse of ONPs and how these rates differ by current use of cigarettes, ENDS, and smokeless tobacco or snus (SLT), as well as sociodemographic characteristics.

## Methods

### Data

We used public-use data from Waves 7 (January 2022 to April 2023, abbreviated as 2022) and 8 (January 2024 to December 2024, abbreviated as 2024) of the PATH Study. Our initial sample included 27,800 adults (ages 18+) who completed Waves 7 and 8. Individuals with missing information on sex, race and ethnicity, age group, ONP use in either Wave, and other tobacco product use in 2022 (including cigarette, ENDS, and SLT use) were excluded. The final sample included 24,640 adults. Race and ethnicity were categorized as non-Hispanic (NH) White, NH Black, or Another or unknown race and ethnicity (with results for this last category suppressed due to non-interpretability). Since few older adults reported ONP use, age group was categorized as 18–24, 25–34, or ≥35 years. Educational attainment was defined as not completed for adults aged <25 years and was categorized as high school or less, some college, bachelor’s degree or more, and not completed or unknown (with results for this last category suppressed due to non-interpretability). Current (established) cigarette use was defined as any past-30-day cigarette use and the lifetime use of at least 100 cigarettes. Current (established) ENDS use was defined as any past-30-day ENDS use and ever “fairly regular” use. Current SLT use was defined as any past-30-day SLT use, including snus pouches, loose snus, moist snuff, dip, spit, and chewing tobacco. SLT use did not include the use of lozenges, gums, or other oral nicotine products. Tobacco product use was categorized five product use states: no current tobacco product use, cigarette-only use, ENDS-only use, SLT-only use, and multiproduct use. Multiproduct use included dual use of cigarettes and ENDS, cigarettes and SLT, ENDS and SLT, and the poly-use of all three products. About 75% of multiproduct use represented dual cigarette and ENDS use. The use of other tobacco products, such as cigars and hookah, was not considered.

To capture ONP use transitions from 2022 to 2024, ONP use at each time point was categorized into never, current, or non-current use in each Wave. Since Wave 7 did not include a question to evaluate regular or established use of ONPs, current use at Waves 7 and 8 was defined as those who used ONPs at least one day in the past 30 days, while non-current use was defined as those who used ONPs at least once in their lifetime but not in the past 30 days. For newly available products like ONPs, surveillance of “ever use, even one time” can overstate real initiation to regular product use. Therefore, current use of ONPs in this analysis does not necessarily imply regular or sustained use. Follow-up time between Waves was approximately two years and was treated as exactly two years in the model. See Table 1 for characteristics of the population by group and ONP use. This analysis of publicly available data was not regulated as human subject research (University of Michigan Institutional Review Board HUM00162265).

**Table 1:** Overall characteristics and prevalence of oral nicotine pouch (ONP) use states in 2022 (Wave 7) and 2024 (Wave 8) among adults in the Population Assessment of Tobacco and Health (PATH) study. Values are weighted percentages (%) and unweighted numbers (N). ENDS = electronic nicotine delivery systems. SLT = smokeless tobacco or snus.

| Group | Overall characteristics |  | ONP use state in 2022 (Wave 7) |  |  |  |  |  | ONP use state in 2024 (Wave 8) |  |  |  |  |  |
| --- | --- | --- | --- | --- | --- | --- | --- | --- | --- | --- | --- | --- | --- | --- |
|  |  |  | Never use |  | Non-current |  | Current use |  | Never use |  | Non-current |  | Current use |  |
|  | % | N | % | N | % | N | % | N | % | N | % | N | % | N |
| Overall | 100 | 24,640 | 96.7 | 23,555 | 2.5 | 802 | 0.8 | 283 | 93.1 | 22,256 | 5.5 | 1,860 | 1.4 | 524 |
| Sex |  |  |  |  |  |  |  |  |  |  |  |  |  |  |
| Female participants | 51.7 | 13,216 | 98.6 | 12,922 | 1.1 | 247 | 0.2 | 47 | 97.0 | 12,536 | 2.7 | 610 | 0.3 | 70 |
| Male participants | 48.3 | 11,424 | 94.6 | 10,633 | 4.0 | 555 | 1.4 | 236 | 89.0 | 9,720 | 8.5 | 1,250 | 2.5 | 454 |
| Race and ethnicity |  |  |  |  |  |  |  |  |  |  |  |  |  |  |
| Non-Hispanic White | 60.4 | 12,857 | 95.7 | 12,047 | 3.3 | 590 | 1.1 | 220 | 91.3 | 11,163 | 6.8 | 1,279 | 1.8 | 415 |
| Non-Hispanic Black | 16.9 | 5,506 | 98.5 | 5,390 | 1.1 | 88 | 0.4 | 28 | 96.1 | 5,174 | 3.3 | 278 | 0.6 | 54 |
| Another or unknown | 22.7 | 6,277 | 97.9 | 6,118 | 1.6 | 124 | 0.4 | 35 | 95.6 | 5,919 | 3.7 | 303 | 0.7 | 55 |
| Age group (years) |  |  |  |  |  |  |  |  |  |  |  |  |  |  |
| 18–24 | 11.7 | 8,188 | 94.6 | 7,833 | 3.9 | 256 | 1.5 | 99 | 86.5 | 7,266 | 9.9 | 682 | 3.6 | 240 |
| 25–34 | 17.5 | 5,781 | 94.7 | 5,454 | 3.8 | 235 | 1.5 | 92 | 89.6 | 5,125 | 7.7 | 495 | 2.6 | 161 |
| ≥35 | 70.8 | 10,671 | 97.5 | 10,268 | 2.0 | 311 | 0.5 | 92 | 95.1 | 9,865 | 4.2 | 683 | 0.7 | 123 |
| Educational attainment |  |  |  |  |  |  |  |  |  |  |  |  |  |  |
| High school or less | 7.9 | 5,491 | 96.7 | 5,238 | 2.7 | 201 | 0.7 | 52 | 93.4 | 4,994 | 5.6 | 431 | 1.0 | 66 |
| Some college | 24.9 | 5,354 | 96.1 | 5,079 | 3.0 | 208 | 0.9 | 67 | 92.8 | 4,815 | 6.0 | 443 | 1.2 | 96 |
| Bachelor's degree or more | 31.6 | 5,547 | 97.9 | 5,348 | 1.5 | 135 | 0.6 | 64 | 95.5 | 5,124 | 3.4 | 301 | 1.1 | 122 |
| Not completed or unknown | 11.9 | 8,248 | 94.6 | 7,890 | 3.9 | 258 | 1.6 | 100 | 86.6 | 7,323 | 9.9 | 685 | 3.5 | 240 |
| Tobacco product use |  |  |  |  |  |  |  |  |  |  |  |  |  |  |
| No other tobacco product use | 81.6 | 18461 | 98.3 | 18033 | 1.3 | 324 | 0.3 | 104 | 96.3 | 17391 | 3.1 | 847 | 0.6 | 223 |
| Cigarette-only use | 10.8 | 3406 | 94.6 | 3229 | 4.7 | 153 | 0.7 | 24 | 88.1 | 3008 | 11.0 | 365 | 1.0 | 33 |
| ENDS-only use | 4.0 | 1660 | 84.6 | 1420 | 11.2 | 178 | 4.1 | 62 | 68.8 | 1155 | 22.3 | 361 | 8.9 | 144 |
| SLT-only use | 1.4 | 350 | 79.6 | 276 | 9.0 | 30 | 11.5 | 44 | 60.7 | 203 | 24.3 | 88 | 14.9 | 59 |
| Multiproduct use | 2.1 | 763 | 77.4 | 597 | 16.8 | 117 | 5.8 | 49 | 64.8 | 499 | 26.3 | 199 | 8.9 | 65 |

### Transition modeling

We used weighted Markov multistate transition models to estimate the underlying transition rates for ONP initiation and cessation in adults overall, and by other tobacco product use and sociodemographic group in univariable models. Although we considered a multivariable model to adjust for multiple exposures, there were too few ONP transitions in certain subpopulations in the dataset to support an adjusted analysis.

Multistate transition models assume that transitions arise from a finite-state stochastic process in which transition hazard rates depend only on the current state, not on past states or transition history (Markov property). The transition hazard rates estimated by the model represent the instantaneous risk of transitioning from one state to another. We specified four allowed transitions between the three ONP use states in the model (Figure 1a): never to non-current use (short-lived experimentation), never to current use (initiation), current use to non-current use (discontinuation), and non-current use to current use (relapse). Although a direct transition from never to non-current use is not possible in theory, inclusion of this transition allowed better alignment of the model estimates with empirical estimates and can be interpreted as short-lived experimentation with ONPs. The associations with this short-lived experimentation may be different than those for initiation to current use.

**Figure 1.**
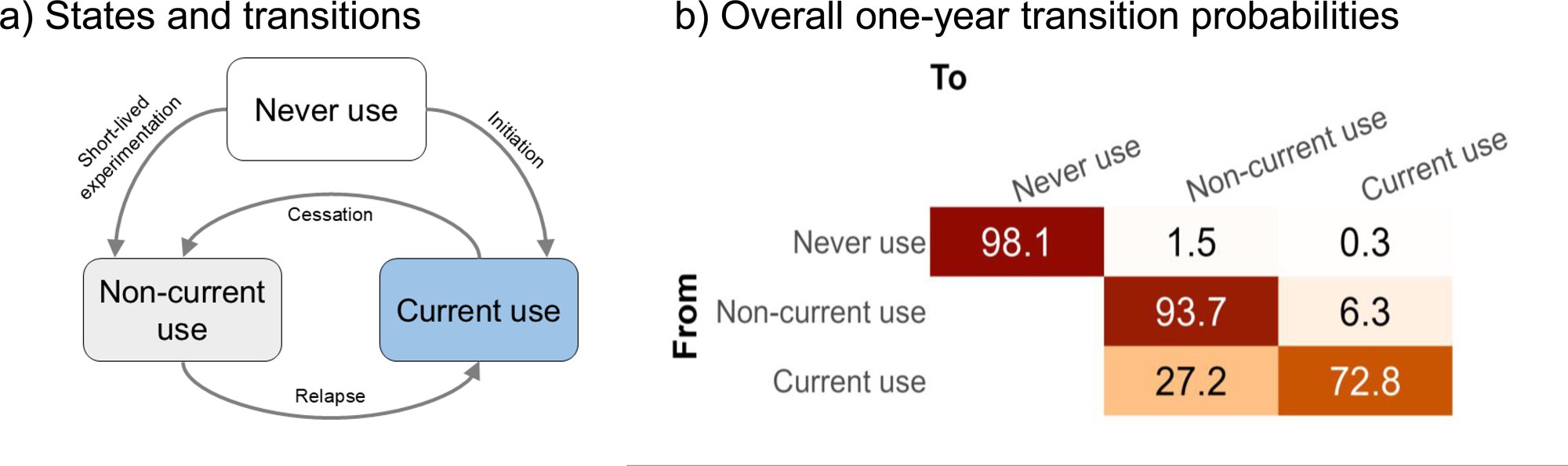
a) Oral nicotine pouch (ONP) use states and direct transitions between states in the model. b) Estimated per-year ONP use transition probabilities in U.S. adults, 2022–24. Confidence intervals are given in eTable 1.

To account for PATH’s complex survey design, we incorporated the PATH Wave 8 single-wave survey weights for the Wave 7 cohort to calculate variances and confidence intervals. Weights were renormalized across ONP use categories, as in Brouwer et al.,^8^ to ensure the representativeness of results within these categories. See eFigure 1 in the eSupplement for a comparison of empirical transition probabilities, estimated transition with the original PATH weights, and estimated transitions with the renormalized weights. Technical details of multistate transition modeling are provided in the eSupplement.

In univariable models, we estimated transition hazard ratios by other tobacco product use and sociodemographic group. We also calculated per-year transition probabilities over the two-year period overall and by population subgroup. All results were estimated using the wmsm function (v.1.4.1)^9^ in R (available at https://tcors.umich.edu/Resources_Research.php), an extension of the msm function^10^ that incorporates complex survey weights.

## Results

In 2022, 0.8% of participants currently used ONPs, 2.5% previously used ONPs (non-current use), and 96.7% had never used ONPs. By 2024, these prevalences changed to 1.4%, 5.5%, and 93.1%, respectively (Table 1). Among adults overall, in one year, we estimated that 1.5% (95% CI: 1.4–1.6%) of those who had never used ONPs would experiment with but discontinue ONP use, and 0.3% (95% CI: 0.3–0.4%) would transition to current use (Figure 1b and eTable 1). We estimated that 27.2% (95% CI: 21.9–32.5%) of those currently using ONPs would discontinue use. For those who used ONPs previously but not currently, 6.3% (95% CI: 5.0–7.6%) were estimated to relapse.

Rates of both ONP experimentation and initiation were higher among participants who used other tobacco products compared to those who used no other tobacco products in 2022 (Figure 2 and eTable 2). Compared to those using no other tobacco products, those using cigarettes-only (HR: 3.40 [95% CI: 2.68–4.32]), ENDS-only (HR: 9.81 [95% CI: 7.74–12.4]), SLT-only (HR: 7.49 [95% CI: 3.92–14.3]), and multiple products (HR: 6.67 [95% CI: 4.25–10.5]) experimented with ONPs at higher rates. Similarly, those using cigarettes-only (HR: 3.34 [95% CI: 1.59–7.02]), ENDS-only (HR: 9.68 [95% CI: 2.89–32.5]), SLT-only (HR: 42.4 [95% CI: 22.3–80.7]), and multiple products (HR: 18.4 [95% CI: 7.11–47.8]) initiated ONP use at higher rates. All product use groups except SLT-only use discontinued ONP use at a higher rate than those using no other tobacco products (cigarette-only HR: 3.02 [95% CI: 1.24–7.34], ENDS-only HR: 2.03 [95% CI: 1.11–3.71], multiproduct use HR: 2.61 [95% CI: 1.12–6.09]). Those using ENDS-only (HR: 6.01 [95% CI: 3.22–11.2]) and multiple products (4.25 [95% CI: 1.75–10.3]) relapsed to ONP use at higher rates than those using no other tobacco products.

**Figure 2.**
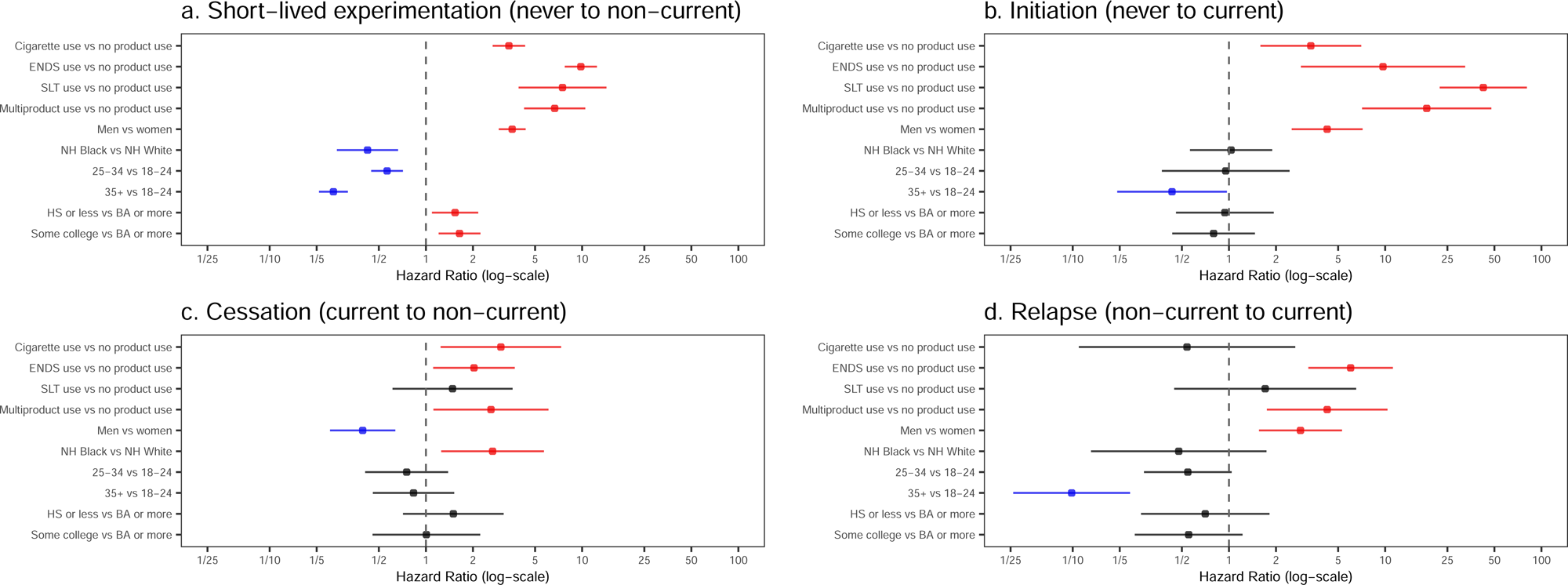
Transition hazard ratios estimated from Waves 7–8 of PATH (2022–24) for a) short-lived experimentation with (never to non-current ONP use) oral nicotine pouch (ONP) use, b) initiation (never to current ONP use), c) cessation (current to non-current ONP use), and d) relapse (non-current to current ONP use) in univariable models by other tobacco product use, sex, race and ethnicity, age group, and educational attainment. Statistically significantly greater hazard ratios are in red, and statistically significantly lower hazard ratios are in blue. The underlying values are given in eTable 2.

In terms of one-year ONP transition probabilities by other tobacco product use, only 0.9% (95% CI: 0.8–1.0%) of those using no other tobacco products were estimated to experiment with ONP use compared to 3.1% (95% CI: 2.5–3.7%) of those using only cigarettes, 7.8% (95% CI: 6.5–9.1%) of those using only ENDS, 7.0% (95% CI: 4.0–9.9%) of those using only SLT, and 6.0% (95% CI: 4.1–7.8%) of those using multiple products (Figure 3 and eTable 1). We estimated that 0.2% (95% CI: 0.1–0.2%) of those using no other tobacco products, 0.4% (95% CI: 0.2–0.6%) of those using only cigarettes, 2.0% (95% CI: 1.0–3.0%) of those using only ENDS, 5.7% (95% CI: 2.7–8.6%) of those using only SLT, and 2.6% (95% CI: 1.0–4.1%) of those using multiple products initiated ONP use. The probability of ONP discontinuation was 18.3% (95% CI: 9.9–26.6%) among those using no other tobacco products, 45.9% (95% CI: 22.0–69.8%) among those using only cigarettes, 30.8% (95% CI: 16.2–45.4%) among those using only ENDS, 25.5% (95% CI: 12.5–38.5%) among those using only SLT, and 38.7% (95% CI: 20.9–56.5%) among those using multiple products. About 3.4% (95% CI: 1.9–4.8%) of those using no other tobacco products, 1.5% (95% CI: 0.0–3.3%) of those using only cigarettes, 16.8% (95% CI: 11.3–22.3%) of those using only ENDS, 5.4% (95% CI: 0.0–11.2%) of those using only SLT, and 11.6% (95% CI: 4.4–18.8%) of those using multiple products relapsed to ONP use.

**Figure 3.**
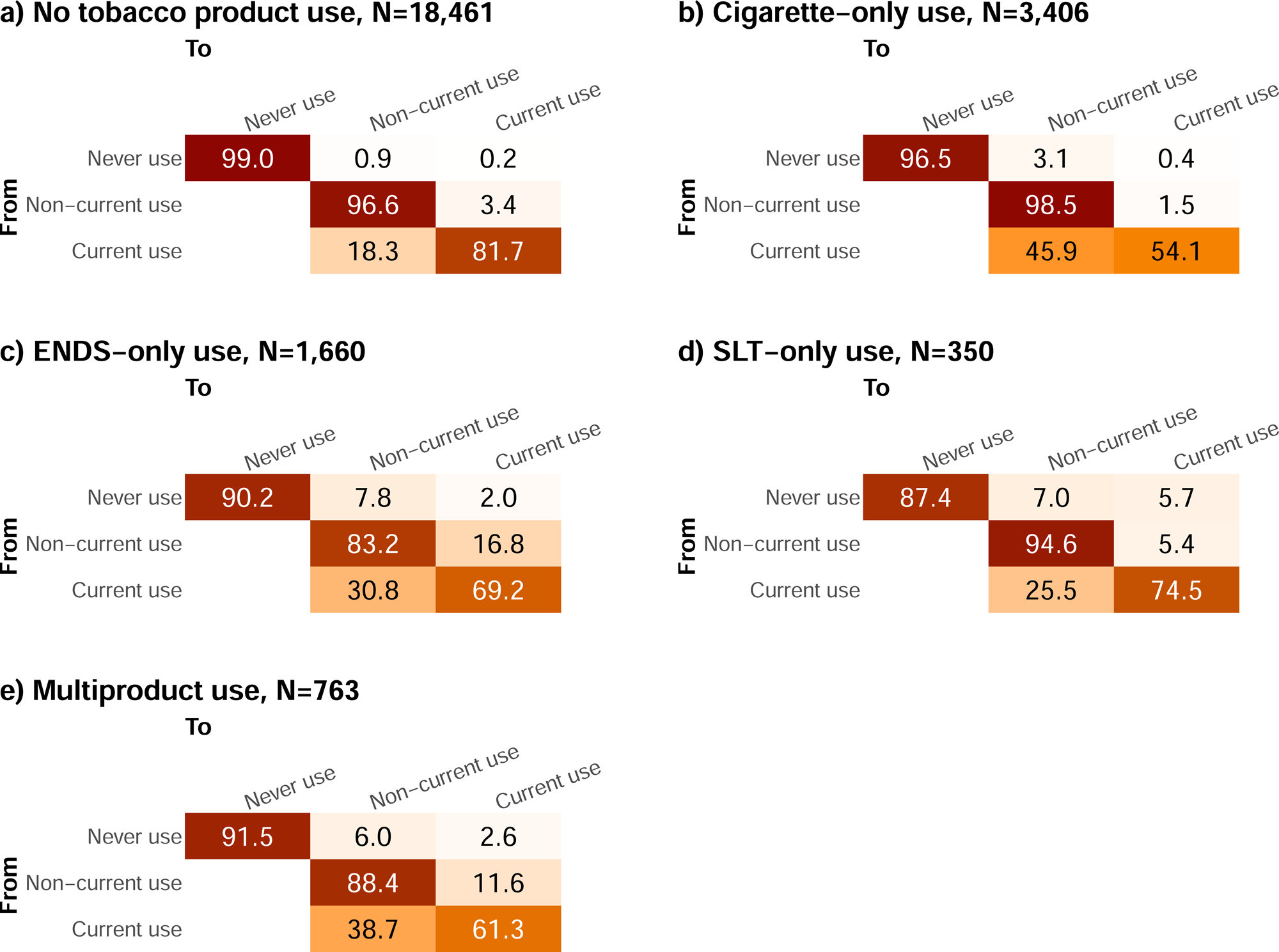
Estimated per-year transition probabilities in oral nicotine pouch (ONP) use among U.S. adults, 2022–24 by other tobacco product use status in 2022: a) no other tobacco product use, b) cigarette-only use, c) ENDS-only use, d) SLT-only use, e) multiproduct use. Confidence intervals are given in eTable 1.

Compared to women, men experimented with (hazard ratio [HR]: 3.57 [95% CI: 2.93–4.35]), initiated (HR: 4.25 [95% CI: 2.52–7.16]), and relapsed to (HR: 2.87 [95% CI: 1.56–5.29]) ONP use at higher rates and discontinued use at a lower rate (HR: 0.39 [95% CI: 0.24–0.64]). In terms of estimated one-year transition probabilities, 2.4% (95% CI: 2.2–2.6%) of men were expected to experiment with ONPs, 0.6% (95% CI: 0.5–0.8%) initiated ONP use, and 7.5% (95% CI: 5.8–9.2%) relapsed to ONP use (Figure 4 and eTable 3). By comparison, only 0.7% (95% CI: 0.6–0.8%) of women experimented with ONPs, 0.1% (95% CI: 0.1–0.1%) initiated ONPs, and 2.2% (95% CI: 1.1–3.3%) relapsed to ONP use. About half (50.4% [95% CI: 35.4–65.3%]) of women using ONPs were expected to discontinue use in one year, compared to less than a quarter (23.5% [95% CI: 18.1–29.0%]) of men.

**Figure 4.**
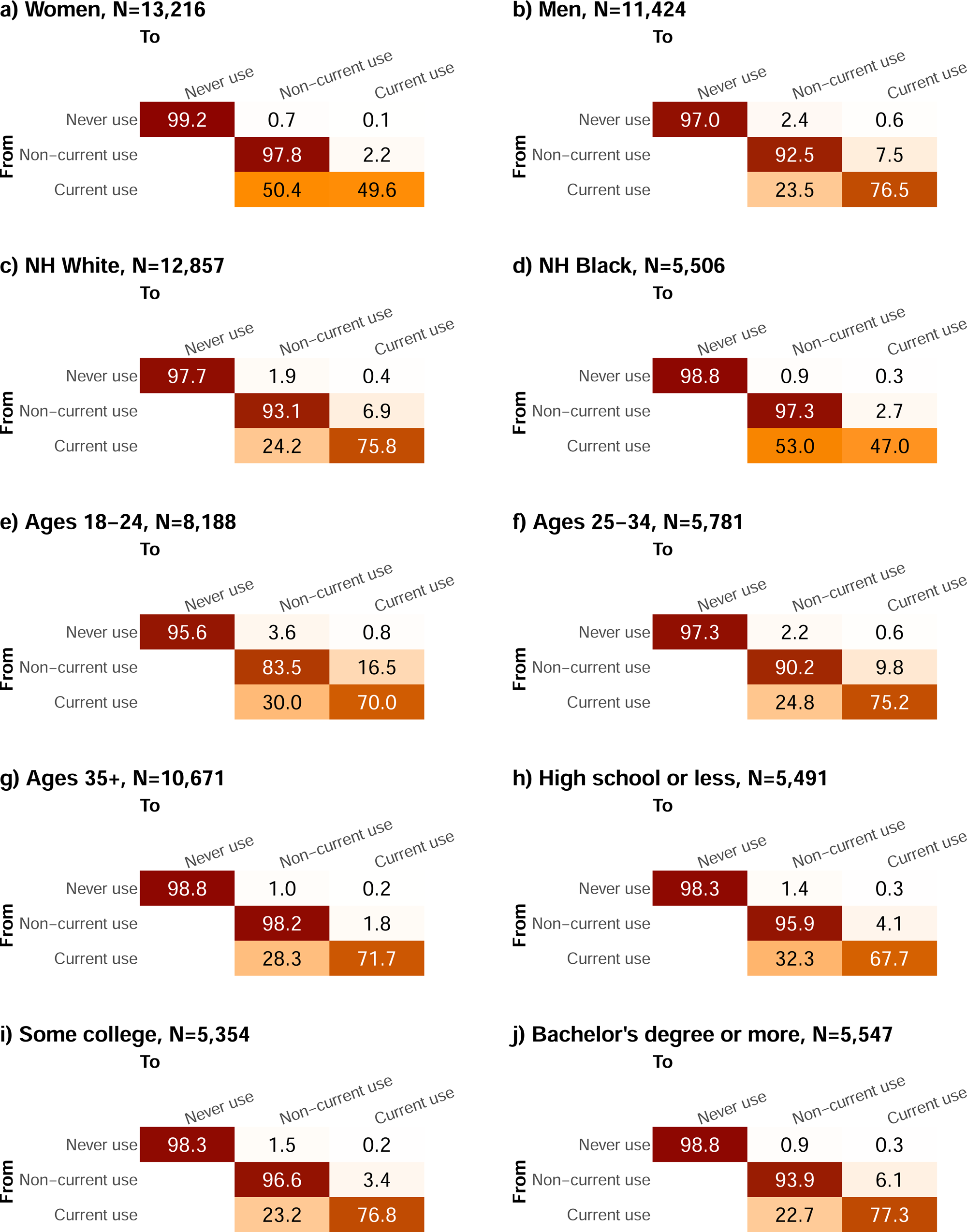
Estimated per-year transition probabilities in oral nicotine pouch (ONP) use among U.S. adults, 2022–24 by sociodemographic characteristics: a) women, b) men, c) non-Hispanic (NH) White, d) NH Black, e) ages 18–24 years, f) ages 25–34 years, g) ages ≥35 years h) high school education or less, i) some college education, and j) bachelor’s degree or more. Confidence intervals are given in eTable 3.

NH Black participants experimented with ONPs at a lower rate than NH White participants (HR: 0.42 [95% CI: 0.27–0.66]) and discontinued current ONP use at a greater rate (HR: 2.67 [95% CI: 1.25–5.70]). Based on estimated transition probabilities, 1.9% (95% CI: 1.7–2.0%) of NH White participants experimented with ONPs in one year, compared with only 0.9% (95% CI: 0.7–1.1%) of NH Black participants. Over half (53.0% [95% CI: 29.3–76.7%]) of NH Black participants using ONPs discontinued use in one year compared to only a quarter (24.2% [95% CI: 18.4–30.0%]) of NH White participants.

Adults ages 25–34 years experimented with (HR: 0.56 [95% CI: 0.45–0.71]) ONP use at lower rates than those ages 18–24 years, while those ages ≥35 years experimented with (0.26 [95% CI: 0.21–0.32]), initiated (HR: 0.43 [95% CI: 0.19–0.97]), and relapsed to (0.10 [95% CI: 0.04–0.23]) ONP use at lower rates than those ages 18–24 years. There were no significant differences in ONP cessation by age group. Translating these results into one-year transition probabilities, 3.6% (95% CI: 3.2–4.0%) of adults ages 18–24 years, 2.2% (95% CI: 1.8–2.5%) of adults ages 25-34 years, and 1.0% (95% CI: 0.9–1.2%) of adults ages ≥35 years experimented with ONPs. An estimated 0.8% (95% CI: 0.6–1.0%) of adults ages 18–24 years and 0.6% (95% CI: 0.4–0.8%) of adults ages 25–34 years initiated ONP use, compared to only 0.2% (95% CI: 0.1–0.3%) of adults ages ≥35 years. An estimated 16.5% (95% CI: 11.6–21.4%) of adults ages 18–24 years, 9.8% (95% CI: 5.8–13.9%) of adults ages 25–34 years, and 1.8% (95% CI: 0.6–3.0%) of adults ages ≥35 years relapsed to ONP use.

Compared to those with a bachelor’s degree or more, those with a high school education or less (HR: 1.54 [95% CI: 1.09–2.16]) or some college education (1.64 [95% CI: 1.21–2.24]) had greater rates of experimenting with ONP use. Across the educational attainment categories, 1.4% (95% CI: 1.1–1.7%) of those with high school education or less, 1.5% (95% CI: 1.2–1.8%) of those with some college education, and 0.9% (95% CI: 0.7–1.1%) of those with a bachelor’s degree or more experimented with ONP use in one year.

## Discussion

We estimated transition rates that underlie the observed transition fractions of short-lived experimentation, initiation, discontinuation, and relapse of ONP use, applying a weighted multistate transition model to a U.S. nationally representative longitudinal cohort, 2022–2024. We found that cigarette-only, ENDS-only, SLT-only, and multiproduct use were all associated with higher rates of both ONP experimentation and initiation compared to those not using these other tobacco products, while those currently using cigarettes-only, ENDS-only, and multiple products, but not necessarily SLT-only, discontinued ONP use at a higher rate. Men experimented with, initiated, and relapsed to ONP use at higher rates and discontinued ONP use at lower rates than women. Those ages 25–34 years experimented with ONP use at lower rates than young adults ages 18–24 years, while those ages ≥35 years experimented with, initiated, and relapsed to ONP use at lower rates than those ages 18–24 years. NH Black participants experimented at a lower rate with and discontinued ONP use at a higher rate than NH White individuals (paralleling similar early associations for ENDS use^11^). Those with less than a high school education and some college experimented with ONPs at a higher rate than those with a bachelor’s degree or more. Given the relatively low prevalence of ONP use in this period, these results were estimated in models that did not account for potential confounding by the other covariates and thus should be interpreted with appropriate caution.

Research on ONPs remains limited given their recent emergence, but previous literature on ONP use supports some of these observed trends. One study found that the prevalence of ever and current ONP use was higher among adults ages 18–34 years, men, and non-Hispanic white individuals compared to their counterparts, and that over half of those who ever used ONPs either currently or formerly smoked cigarettes.^12^ Among tobacco-naïve adults, ever and current use of ONPs is rare, with increased odds of ever use of ONPs for those currently using SLT, cigarettes, and e-cigarettes.^13^

An important finding from our analysis is that those using only cigarettes, ENDS, or multiple tobacco products were, in this period, more likely than those who did not use any products to transition *both* into and out of current ONP use. This result is not unexpected, given the relatively recent emergence of ONPs in the market and that higher lifetime ONP use has been found among those using other tobacco products.^14^ Cigarette manufacturers have also targeted ONP advertisements towards cigarette users as an alternative for when they are in a situation where they can’t smoke.^15^ This marketing may have contributed to ONP experimentation among those using other tobacco products during this period. There is also some evidence to support that interest in using ONPs is associated with plans to quit cigarette smoking or a prior cigarette quit attempt, and that current and daily use of pouches is highest among adults who have recently quit another tobacco product.^13,16^ However, these early findings do not necessarily imply long-term cessation of other products or sustained use of ONPs.

Although it is expected that ONPs will be found to be considerably less harmful than cigarettes, labels such as “tobacco-free” and “flavor ban approved” on many ONP products may lead consumers to believe these products pose little risk, and there remains discussion about whether the FDA should allow them.^17^ In particular, a study found that the “non-tobacco” warning label was associated with being more likely to perceive ONPs as less harmful than SLT products.^18^ In June of 2026, the FDA authorized 20 Zyn nicotine products to be marketed with the modified risk claim, “Using Zyn instead of cigarettes puts you at a lower risk of mouth cancer, heart disease, lung cancer, stroke, emphysema, and chronic bronchitis.”^19^ These labels may lead those using other tobacco products to switch—or at least experiment with trying to switch—to ONPs. However, limitations of the current analysis are that our transition model only includes states for ONP use, with other tobacco products included as covariates instead, and that PATH is an observational study. Therefore, this analysis does not necessarily address the question that tobacco product users are replacing cigarettes or other tobacco products with ONPs. We will investigate this in future work.

Although this analysis found preliminary associations between cigarette, ENDS, SLT, and multiproduct use and ONP initiation in 2022–24, continued transition surveillance and more research are needed to determine whether ONP use among other product users might remain primarily a transient experimental state, replace other tobacco product use, or lead to sustained poly-product use. Higher ONP cessation rates among those using cigarettes, ENDS, and multiple products may suggest ONP experimentation rather than sustained use in the 2022–24 period covered by the analysis, but, as with ENDS previously, these transitions may evolve over time. Additionally, differences in ONP initiation and cessation rates by sex, race and ethnicity, age group, and educational attainment point to differences in susceptibility to ONP use by sociodemographic group. Continued monitoring of ONP use transitions over time will be necessary to continuously inform tobacco control efforts as these products become more established.

## Supporting information

eSupplement

## Data Availability

Public Use Files from the Population Assessment of Tobacco and Health (PATH) Study are available for download from an open access repository (https://doi.org/10.3886/ICPSR36498.v25). Conditions of use are available on the aforementioned website.

## Acknowledgments

This project was funded through National Cancer Institute (NCI) and Food and Drug Administration (FDA) grant U54CA229974. The opinions expressed in this article are the authors’ own and do not necessarily reflect the views of the National Institutes of Health, the Department of Health and Human Services, or the United States government.

## Author Contributions

Conceptualization AFB, LMS-R, DTL, RMe; methodology: AFB; data curation: JJ and EJ-M; analysis and original draft preparation: OKR; review and editing: OKR, LMS-R, EJ-M, JJ, DTL, RMe, and AFB; funding acquisition: RMe and DTL; guarantor: AFB.

## Data Sharing Statement

Public Use Files from the Population Assessment of Tobacco and Health (PATH) Study are available for download from an open access repository (https://doi.org/10.3886/ICPSR36498.v25).^20^ Conditions of use are available on the aforementioned websites.

