## Supplementary material for "Rates of oral nicotine pouch experimentation, initiation, and cessation by other tobacco product use and sociodemographic characteristics among U.S. adults, 2022–2024": eSupplement

**Multistate transition modeling**

This technical appendix is reproduced in part from the supplementary material of Brouwer et al^1^ but includes updated notation and technical details. Additional details may be found in Jackson^2^, and a tutorial for working with the weighted multistate transition model (wmsm) code is available at <https://tcors.umich.edu/Resources_Research.php> along with the code itself.

A multistate transition model is a continuous-time, finite-state stochastic process with the Markov assumption that transition rates depend only on the current state and not on past states or transition history. We denote the state of the process at time $t$ as $S\left( t \right)$. We denote the probability that an individual is in state $j$ after an amount of $\Delta t$ since they were observed in state $i$ as

$$\begin{aligned} P_{ij}(t,t+\Delta t)=Pr\left[ S\left( t+\Delta t \right)=j | S\left( t \right)=i \right]. \#\left( 1 \right) \end{aligned}$$

In general, the transition probabilities can depend on the observation time $t$ in addition to the time span $\Delta t$, and in this analysis we consider discrete time periods over which we assume that that the model is homogeneous in time, i.e., $P_{ij}\left( t,t+\Delta t \right)= P_{ij}(0,\Delta t)$. Thus, we drop the dependence on $t$ moving forward and write $P_{ij}(\Delta t)$. We then define the hazard rate of the transition from state $i$ to state$j$, for $i\neq j$, as

$$\begin{aligned} q_{ij}=\lim_{\Delta t\to0} \frac{1}{\Delta t}Pr\left[ S\left( \Delta t \right)=j | S\left( 0 \right)=i \right]. \#\left( 2 \right) \end{aligned}$$

Transition hazards ratios $\rho_{ij,c_{l}}$may be determined for each transition $i$ to $j$ for each level $l$ of a covariate $c$, relative to the referent $c_{0}$: $q_{ij,c_{l}}=\rho_{ij,c_{l}}\times q_{ij,c_{0}}$. For time-varying covariates, a participant’s transition rate is determined by their characteristic at their most recent previous observation.

The transition hazard rates form a matrix $Q = \left[ q_{ij} \right],$ where the diagonal entries are given by $q_{ii}=-\sum_{j\neq i} q_{ij}$. The transition probability matrix $P\left( \Delta t \right)=\left[ P_{ij}\left( \Delta t \right) \right]$ is a function of the transition hazard rates and may be calculated as the matrix exponential of $\Delta t\cdot Q$, that is

$\begin{aligned} P\left( \Delta t \right)=\text{expm}\left( \Delta t\cdot Q \right).\#\left( 3 \right) \end{aligned}$

Transition probabilities can be estimate for any value of $\Delta t$, assuming that the transition hazards $q_{ij}$ do not change over that time period (assumption of homogeneity).

The values of the transition hazard rates $q_{ij}$of multistate transition model are estimated by maximizing a statistical likelihood $L$ given a set of observed states and times by comparing the observed states to the probabilities in $P\left( \Delta t \right)$ as a function of the transition hazard matrix $Q$. Specifically, consider a set of individuals $m = 1, \ldots,N$ and their observed states $s_{m,t_{m,k}}$at times $t_{m,k}$, where $k$ is the index of individual $m$’s $K_{m}$observations in the data. Denote the data as $s=\{s_{m,t_{m,k}}\}$. We assume the individuals are independent, and thus we multiply all the modeled probabilities of the observed transitions:

$$\begin{aligned} L\left( Q|s \right)=\prod_{m=1}^{N} \prod_{k=1}^{K_{m}-1} P_{s_{m,t_{m,k}},s_{m,t_{m,k+1}}}(t_{m,k+1}-t_{m,k}).\#\left( 4 \right) \end{aligned}$$

where $P_{i,j}$ is a function of $Q$ as in Eqn (3).

Participant weights $W_{m}$can be incorporated into a weighted likelihood $L^{*}$. Although it is not strictly necessary to normalize the weights to the population size, $w_{m}=N\cdot W_{m}/\sum_{v} W_{v}$, it is convenient to do so because the resulting likelihood will correspond to the unweighted likelihood when all participants have equal weight. The weighted likelihood is given by

$$\begin{aligned} L^{*}\left( Q|s \right)=\prod_{m=1}^{N} \prod_{k=1}^{K_{m}-1} \left( P_{s_{m,t_{m,k}},s_{m,t_{m,k+1}}}(t_{m,k+1}-t_{m,k}) \right)^{w_{m}}.\#\left( 5 \right) \end{aligned}$$

Following the msm package^2^, we assume that estimated transition hazard rates $\hat{q}_{ij}$ are normally distributed on the log-scale, that is

$$\begin{aligned} \log\hat{q}_{ij}\sim N\left( \mu=\text{mean}\left( \log\hat{q}_{ij} \right),\sigma^{2}=V\left( \log\hat{q}_{ij} \right) \right),\#\left( 6 \right) \end{aligned}$$

where $V$ denotes the estimated variance.

We estimated weighted point estimates for the log transition hazard rates $\log\hat{q}_{ij}$ by minimizing ${-logL}^{*}\left( Q|s \right)$as a function of the transition hazard rates. (This approach is equivalent to maximum likelihood estimation).

Variance estimates $V\left( \log\hat{q}_{ij} \right)$are calculated using replicate weights $w_{m}^{r}$. Replicate weights are a way to account for complex survey design aspects, such as strata and primary sampling units. PATH uses a variant of balanced repeated replication called Fay’s method to calculate 100 replicate weights. We calculate $\log\hat{q}_{ij}^{r}$ for each $r$ as above. Then, we calculate the variance of $\log\hat{q}_{ij}^{r}$ as

$$\begin{aligned} V\left( \log\hat{q}_{ij} \right)=c\sum_{r=1}^{100} \left( \log\hat{q}_{ij}^{r}-\log\hat{q}_{ij} \right)^{2}\#\left( 7 \right) \end{aligned}$$

where $c=1/(100\left( 1-0.3 \right)^{2})$ as specified by PATH.

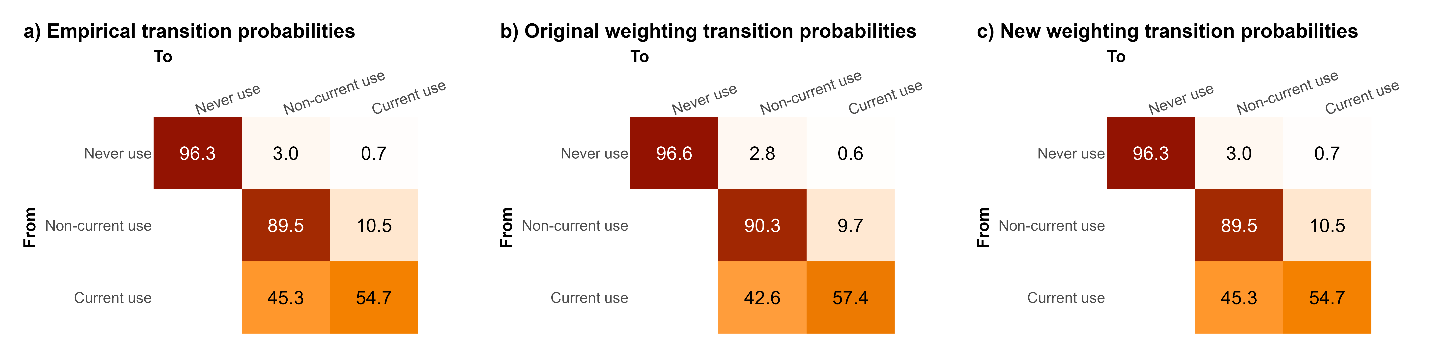

**eFigure 1.** a) Empirical two-year transition probabilities for all adults in PATH Waves 7–8 (2022–24), b) Modeled two-year transition probabilities using the original PATH Wave 8 all-waves survey weights for the Wave 7 cohort, c) Modeled two-year transition probabilities using the PATH Wave 8 all-waves survey weights for the Wave 7 cohort renormalized so that each product use state has equal weight. The renormalized weights resulted in a better representation of the empirical results stratified by ONP product use.

| ONP Transition | Overall  %  (95% CI) | No tobacco product use  %  (95% CI) | Cigarette-only use  %  (95% CI) | ENDS-only use  %  (95% CI) | SLT-only use  %  (95% CI) | Multiproduct use  %  (95% CI) |
| --- | --- | --- | --- | --- | --- | --- |
| Never use persistence | 98.1  (98.0, 98.3) | 99.0  (98.8, 99.1) | 96.5  (95.9, 97.1) | 90.2  (88.7, 91.7) | 87.4  (84.2, 90.6) | 91.5  (89.2, 93.7) |
| Never use to non-current use | 1.5  (1.4, 1.6) | 0.9  (0.8, 1.0) | 3.1  (2.5, 3.7) | 7.8  (6.5, 9.1) | 7.0  (4.0, 9.9) | 6.0  (4.1, 7.8) |
| Never use to current use | 0.3  (0.3, 0.4) | 0.2  (0.1, 0.2) | 0.4  (0.2, 0.6) | 2.0  (1.0, 3.0) | 5.7  (2.7, 8.6) | 2.6  (1.0, 4.1) |
| Non-current use to never use | — | — | — | — | — | — |
| Non-current use persistence | 93.7  (92.4, 95.0) | 96.6  (95.2, 98.1) | 98.5  (96.7, 100.0) | 83.2  (77.7, 88.7) | 94.6  (88.8, 100.0) | 88.4  (81.2, 95.6) |
| Non-current use to current use | 6.3  (5.0, 7.6) | 3.4  (1.9, 4.8) | 1.5  (0.0, 3.3) | 16.8  (11.3, 22.3) | 5.4  (0.0, 11.2) | 11.6  (4.4, 18.8) |
| Current use to never use | — | — | — | — | — | — |
| Current use to non-current use | 27.2  (21.9, 32.5) | 18.3  (9.9, 26.6) | 45.9  (22.0, 69.8) | 30.8  (16.2, 45.4) | 25.5  (12.5, 38.5) | 38.7  (20.9, 56.5) |
| Current use persistence | 72.8  (67.5, 78.1) | 81.7  (73.4, 90.1) | 54.1  (30.2, 78.0) | 69.2  (54.6, 83.8) | 74.5  (61.5, 87.5) | 61.3  (43.5, 79.1) |

**eTable 1.** One-year oral nicotine pouch (ONP) transition probabilities and 95% confidence intervals estimated from Waves 7–8 of PATH (2022–24) overall and by other tobacco product use: no other tobacco product use, cigarette-only use, electronic nicotine delivery system only (ENDS-only) use, smokeless tobacco and snus only (SLT-only) use, and multiproduct use.

**eTable 2.** Transition hazard ratios and 95% confidence intervals for oral nicotine pouch (ONP) use transitions from Waves 7–8 of PATH (2022–24) in unadjusted models by other tobacco product use, sex, race and ethnicity, age group, and educational attainment. ENDS = electronic nicotine delivery system. SLT = smokeless tobacco and snus.

|  | Never use to non-current use | | Never use to current use | | Non-current use to current use | | Current use to non-current use | |
| --- | --- | --- | --- | --- | --- | --- | --- | --- |
|  | Estimate | 95% CI | Estimate | 95% CI | Estimate | 95% CI | Estimate | 95% CI |
| Tobacco product use |  |  |  |  |  |  |  |  |
| Cigarette-only use vs no current use | **3.40** | **(2.68, 4.32)** | **3.34** | **(1.59, 7.02)** | 0.54 | (0.11, 2.66) | **3.02** | **(1.24, 7.34)** |
| ENDS-only use vs no current use | **9.81** | **(7.74, 12.4)** | **9.68** | **(2.89, 32.5)** | **6.01** | **(3.22, 11.2)** | **2.03** | **(1.11, 3.71)** |
| SLT-only use vs no current use | **7.49** | **(3.92, 14.3)** | **42.4** | **(22.3, 80.7)** | 1.71 | (0.45, 6.52) | 1.48 | (0.61, 3.59) |
| Multiproduct use vs no current use | **6.67** | **(4.25, 10.5)** | **18.4** | **(7.11, 47.8)** | **4.25** | **(1.75, 10.3)** | **2.61** | **(1.12, 6.09)** |
| Sex |  |  |  |  |  |  |  |  |
| Men vs women | **3.57** | **(2.93, 4.35)** | **4.25** | **(2.52, 7.16)** | **2.87** | **(1.56, 5.29)** | **0.39** | **(0.24, 0.64)** |
| Race & ethnicity |  |  |  |  |  |  |  |  |
| Non-Hispanic Black vs Non-Hispanic White | **0.42** | **(0.27, 0.66)** | 1.03 | (0.56, 1.89) | 0.48 | (0.13, 1.74) | **2.67** | **(1.25, 5.70)** |
| Age group (years) |  |  |  |  |  |  |  |  |
| 25*–*34 vs 18*–*24 | **0.56** | **(0.45, 0.71)** | 0.95 | (0.37, 2.44) | 0.55 | (0.29, 1.04) | 0.75 | (0.41, 1.39) |
| ≥35 vs 18*–*24 | **0.26** | **(0.21, 0.32)** | **0.43** | **(0.19, 0.97)** | **0.10** | **(0.04, 0.23)** | 0.83 | (0.46, 1.52) |
| Education |  |  |  |  |  |  |  |  |
| High school or less vs bachelor’s or more | **1.54** | **(1.09, 2.16)** | 0.94 | (0.46, 1.93) | 0.71 | (0.27, 1.82) | 1.50 | (0.71, 3.15) |
| Some college vs bachelor’s or more | **1.64** | **(1.21, 2.24)** | 0.80 | (0.43, 1.47) | 0.55 | (0.25, 1.22) | 1.01 | (0.46, 2.23) |

| ONP Transition | Women  %  (95% CI) | Men  %  (95% CI) | Non-Hispanic White  %  (95% CI) | Non-Hispanic Black  %  (95% CI) | Ages 18–24  %  (95% CI) | Ages 25–34  %  (95% CI) | Ages ≥35  %  (95% CI) | High school or less  %  (95% CI) | Some college  %  (95% CI) | Bachelor’s degree or more  %  (95% CI) |
| --- | --- | --- | --- | --- | --- | --- | --- | --- | --- | --- |
| Never use persistence | 99.2  (99.1, 99.3) | 97.0  (96.8, 97.2) | 97.7  (97.5, 97.9) | 98.8  (98.5, 99.0) | 95.6  (95.2, 96.0) | 97.3  (96.9, 97.7) | 98.8  (98.6, 98.9) | 98.3  (98.0, 98.6) | 98.3  (98.0, 98.5) | 98.8  (98.6, 99.0) |
| Never use to non-current use | 0.7  (0.6, 0.8) | 2.4  (2.2, 2.6) | 1.9  (1.7, 2.0) | 0.9  (0.7, 1.1) | 3.6  (3.2, 4.0) | 2.2  (1.8, 2.5) | 1.0  (0.9, 1.2) | 1.4  (1.1, 1.7) | 1.5  (1.2, 1.8) | 0.9  (0.7, 1.1) |
| Never use to current use | 0.1  (0.1, 0.1) | 0.6  (0.5, 0.8) | 0.4  (0.3, 0.5) | 0.3  (0.2, 0.4) | 0.8  (0.6, 1.0) | 0.6  (0.4, 0.8) | 0.2  (0.1, 0.3) | 0.3  (0.2, 0.4) | 0.2  (0.1, 0.3) | 0.3  (0.2, 0.4) |
| Non-current use to never use | — | — | — | — | — | — | — | — | — | — |
| Non-current use persistence | 97.8  (96.7, 98.9) | 92.5  (90.8, 94.2) | 93.1  (91.5, 94.7) | 97.3  (94.4, 100.) | 83.5  (78.6, 88.4) | 90.2  (86.1, 94.2) | 98.2  (97.0, 99.4) | 95.9  (93.1, 98.7) | 96.6  (94.5, 98.6) | 93.9  (90.9, 96.8) |
| Non-current use to current use | 2.2  (1.1, 3.3) | 7.5  (5.8, 9.2) | 6.9  (5.3, 8.5) | 2.7  (0.0, 5.6) | 16.5  (11.6, 21.4) | 9.8  (5.8, 13.9) | 1.8  (0.6, 3.0) | 4.1  (1.3, 6.9) | 3.4  (1.4, 5.5) | 6.1  (3.2, 9.1) |
| Current use to never use | — | — | — | — | — | — | — | — | — | — |
| Current use to non-current use | 50.4  (35.4, 65.3) | 23.5  (18.1, 29.0) | 24.2  (18.4, 30.0) | 53.0  (29.3, 76.7) | 30.0  (19.7, 40.4) | 24.8  (15.2, 34.4) | 28.3  (20.4, 36.2) | 32.3  (19.7, 44.8) | 23.2  (12.7, 33.7) | 22.7  (11.8, 33.5) |
| Current use persistence | 49.6  (34.7, 64.6) | 76.5  (71.0, 81.9) | 75.8  (70.0, 81.6) | 47.0  (23.3, 70.7) | 70.0  (59.6, 80.3) | 75.2  (65.6, 84.8) | 71.7  (63.8, 79.6) | 67.7  (55.2, 80.3) | 76.8  (66.3, 87.3) | 77.3  (66.5, 88.2) |

**eTable 3.** One-year oral nicotine pouch (ONP) transition probabilities and 95% confidence intervals estimated from Waves 7–8 of PATH (2022–24) by sociodemographic characteristics: women, men, non-Hispanic White, Non-Hispanic Black, 18–24, 25–34, ≥35, high school or less, some college, and bachelor’s degree or more.
